# Building Up Facilitators, Breaking Down Barriers: A Scoping Review Mapping Factors that Impact Participation in Vaccine Trials

**DOI:** 10.1101/2025.08.21.25331718

**Authors:** Toni Claessens, Aurélie De Waele, Greet Hendrickx, Margot Hellemans, Laura Willen, Pierre Van Damme

**Affiliations:** University of Antwerp, Antwerp, Belgium

**Keywords:** vaccine trials, trial participation, COVID-19, vaccine hesitancy, barriers and facilitators, underrepresented groups, scoping review, health communication

## Abstract

**Background:** Vaccine trials are critical for developing and approving vaccines, yet ensuring representative participation remains challenging. The COVID-19 pandemic underscored both the importance of vaccination and the pervasive issue of vaccine hesitancy, a psychological state of indecisiveness when making a vaccine-related decision. This hesitancy extends to vaccine trials, which are essential for generating reliable data on safety, efficacy, and potential side effects. The present scoping review identifies key barriers and facilitators influencing trial participation, focusing on COVID-19 trials for the general population and broader vaccine trials for three underrepresented groups (pregnant women, older adults, and parents).

**Methods:** We conducted systematic PubMed searches (April 1–June 20, 2022) using MeSH terms and keywords to locate relevant biomedical literature on COVID-19 vaccine trial participation. Of 2,924 unique articles, 193 were screened in full, and 56 met inclusion criteria.

**Results:** Safety concerns, mistrust, and lack of information emerged as primary barriers to vaccine trial enrolment. Conversely, altruism, trust in researchers and vaccines, and incentives, especially medical incentives, were identified as facilitators. Understanding these factors is critical for expanding volunteer registries and improving pandemic preparedness.

**Conclusion:** Addressing social, behavioural, and practical determinants of vaccine trial participation can foster greater enrolment and more robust data collection. Insights from this review offer guidance for health authorities aiming to enhance communication strategies and encourage broader participation in vaccine trials, ultimately supporting global efforts to control infectious diseases.

## Introduction

Following the World Health Organization’s (WHO) declaration of COVID-19 as a pandemic in March 2020, the quest for effective COVID-19 vaccines became a global priority, in addition to ongoing efforts for vaccines against other diseases. The approval of the first safe and effective COVID-19 vaccine by the European Commission in December 2020 marked a significant milestone, prompting widespread vaccination campaigns. While these efforts were crucial in controlling the pandemic, they depended on a cornerstone of vaccine development: robust participation in vaccine trials. Despite the clear importance and urgency of developing effective vaccines amidst the pandemic, securing sufficient participation in clinical trials remained a major challenge (Cobb et al., 2014; Detoc et al., 2019; Swan et al., 2009).

Vaccine trials are a crucial component in the development and approval of vaccines, providing essential data to ensure vaccines are both safe and effective, yet recruitment remains a persistent challenge in various types of clinical trial management (Harrington et al., 2017). However, while significant research exists on vaccine hesitancy and the willingness of populations to be vaccinated (De Figueiredo et al., 2022; De Figueiredo et al., 2020; Larson et al., 2019; Steinert et al., 2022), far less is known about the participation in vaccine trials specifically. This knowledge gap is particularly concerning, as low participation can delay vaccine development and hinder public health responses. Compared to other types of clinical studies, participation in vaccine trials presents distinct challenges. A nationally representative survey conducted in the United States found that the public is less likely to participate in vaccine trials than in studies for new medications or medical devices, suggesting that vaccine trials face specific recruitment barriers (Cobb et al., 2014). Beyond common challenges of clinical trial recruitment, such as mistrust in research or difficulties in identifying and retaining participants (McDonald et al., 2006; Tramm et al., 2013), vaccine trials are further complicated by concerns and misconceptions specific to vaccines, many of which remain insufficiently explored (Detoc et al., 2017). Therefore, identifying and addressing the factors that affect recruitment is essential to help researchers meet their targets and ensure timely progress in vaccine development (Harrington et al., 2017).

While studies examining factors influencing participation in vaccine trials exist, the literature remains fragmented. Findings are often embedded within broader studies rather than being the primary focus, spanning multiple countries, utilising diverse research methodologies, and investigating a wide range of variables. This fragmentation has made it challenging to derive a comprehensive overview of the factors influencing vaccine trial participation. However, a recent narrative review by Dean et al. (2023) provides a first structured synthesis of barriers and facilitators to vaccine trial participation, addressing this gap to some extent. Their review highlights a wide range of factors but primarily focuses on vaccine trials in general.

We argue, however, that the COVID-19 pandemic may have fundamentally altered the landscape of vaccine trial participation. It may have heightened pre-existing factors or introduced entirely new ones, prompting the idea to focus specifically on factors associated with participation in COVID-19 vaccine trials. During the COVID-19 vaccination campaign, a disperse uptake of vaccines was reported over different countries, revealing hesitancy towards these newly introduced vaccines (Berry et al., 2021; De Figueiredo & Larson, 2021; Ditekemena et al., 2021; Freeman et al., 2020; Head et al., 2020; Steinert et al., 2022). A scoping review on the relationship between the pandemic and vaccine hesitancy showed that common factors contributing to hesitancy towards COVID-19 vaccines included concerns about vaccine safety, and their effectiveness (de Albuquerque Veloso Machado et al., 2021). The rapid development and implementation of COVID-19 vaccines fuelled these perceptions of potentially higher risks of side effects, putting the public’s trust to the test (Lin et al., 2020). Another factor associated with an increased fear of side effects was the circulation of misinformation on COVID-19 vaccines (de Albuquerque Veloso Machado et al., 2021), paving the way to what has been termed an ‘infodemic’ (Islam et al., 2020). At the same time, mistrust of health authorities has been shown to affect vaccine readiness, but also willingness to participate in vaccine trials (Strauss et al., 2001). Given the overlap between vaccine hesitancy and vaccine trial participation, these concerns may have similarly impacted the willingness to enrol in COVID-19 vaccine trials (Detoc et al., 2019).

This hesitancy or unwillingness to participate in vaccine trials might be even more pronounced among underrepresented groups - including racial minorities, older adults, people with disabilities, immunocompromised individuals, pregnant women and children and adolescents - who are often excluded from initial trials (Corbie-Smith et al., 2002; Flores et al., 2021). The WHO emphasized in their 152nd executive board session that “clinical trials are likely to produce the clearest result when performed in different settings, including all major population groups, with a particular focus on underrepresented populations”. Ensuring their inclusion is not only a matter of equity but also of public health necessity. For example, vaccine trials involving pregnant women help determine safety and dosage guidelines, directly shaping vaccination policies for this population. In a pandemic situation, which disproportionately affects vulnerable populations, the success of these trials hinges upon the active participation of diverse demographic groups (Mackey et al., 2021; Tai et al., 2021). However, scaling up representation in vaccine trials goes beyond simply expanding eligibility criteria. It requires efforts to address the unique barriers that prevent these groups from participation in vaccine trials(Corbie-Smith et al., 2002). While sufficient research exists on COVID-19 vaccine trials for the general population, comparatively little COVID-19 research focused on underrepresented groups, a gap that was also evident in the review from Dean et al. (2023), which did not specifically consider vulnerable populations. Therefore, this review broadens its scope beyond COVID-19 and will draw on literature from trials involving a variety of diseases and vaccines, offering a better understanding of factors affecting vaccine trial participation for these groups. This approach allows us to address the gap in COVID-19-specific data for vulnerable populations while ensuring their inclusion in future vaccine development efforts.

To address gaps in the literature, this review builds upon prior research by focusing specifically on COVID-19 vaccine trial participation and on studies that examine the experiences of vulnerable groups, both of which remain underexplored. By integrating these dimensions, it provides a more targeted and timely analysis of the evolving landscape of vaccine trial participation. Overall, this manuscript synthesises existing research on the barriers and facilitators of vaccine trial participation, offering valuable insights for improving recruitment strategies and fostering more inclusive vaccine development efforts.

## Method

We conducted a scoping review to systematically examine the existing literature on participation in (COVID-19) vaccine trials. A scoping review offers a comprehensive approach combining diverse sources of evidence, providing a broad overview of the landscape of research on the topic (Arksey & O’Malley, 2005). Our methodology followed established guidelines. We conducted a series of PUBMED searches – designed using Medical Subject Headings (MeSH) terms and keywords – to identify relevant biomedical literature on COVID-19 vaccine trial participation from Medline, life science journals and online books. Searches were conducted between April 1^st^, 2022, and June 20^th^, 2022. The PUBMED-search strategies used to underpin this scoping review will be described further and are depicted in *Figure 1*. The following inclusion criteria applied: peer-reviewed empirical research (quantitative or qualitative), full-text article available in English, and addressing (a combination of) the following keywords: *vaccine trial participation*, *COVID-19*, *vaccine hesitancy*, *pregnant women*, *elderly*, and/or *children and adolescents*. Searching literature was conducted to identify the barriers and facilitators for participation in COVID-19 vaccine trials. Additionally, three subgroups known to be currently underrepresented in vaccine trials were selected - pregnant women, children and adolescents, and older adults - and their barriers and facilitators for participation in vaccine trials in general were identified (including non-COVID-19 vaccine trials). We have chosen to use the term ‘older adults’ instead of our original search term ‘elderly’ as it is considered more respectful and inclusive. The term ‘elderly’ often carries connotations of being ‘very old’ and can exclude individuals who are in the earlier stages of aging. Studies conducted both within and outside the EU were included in this scoping review. The full list of included articles can be found in *Table 1*.

**Figure 1:**
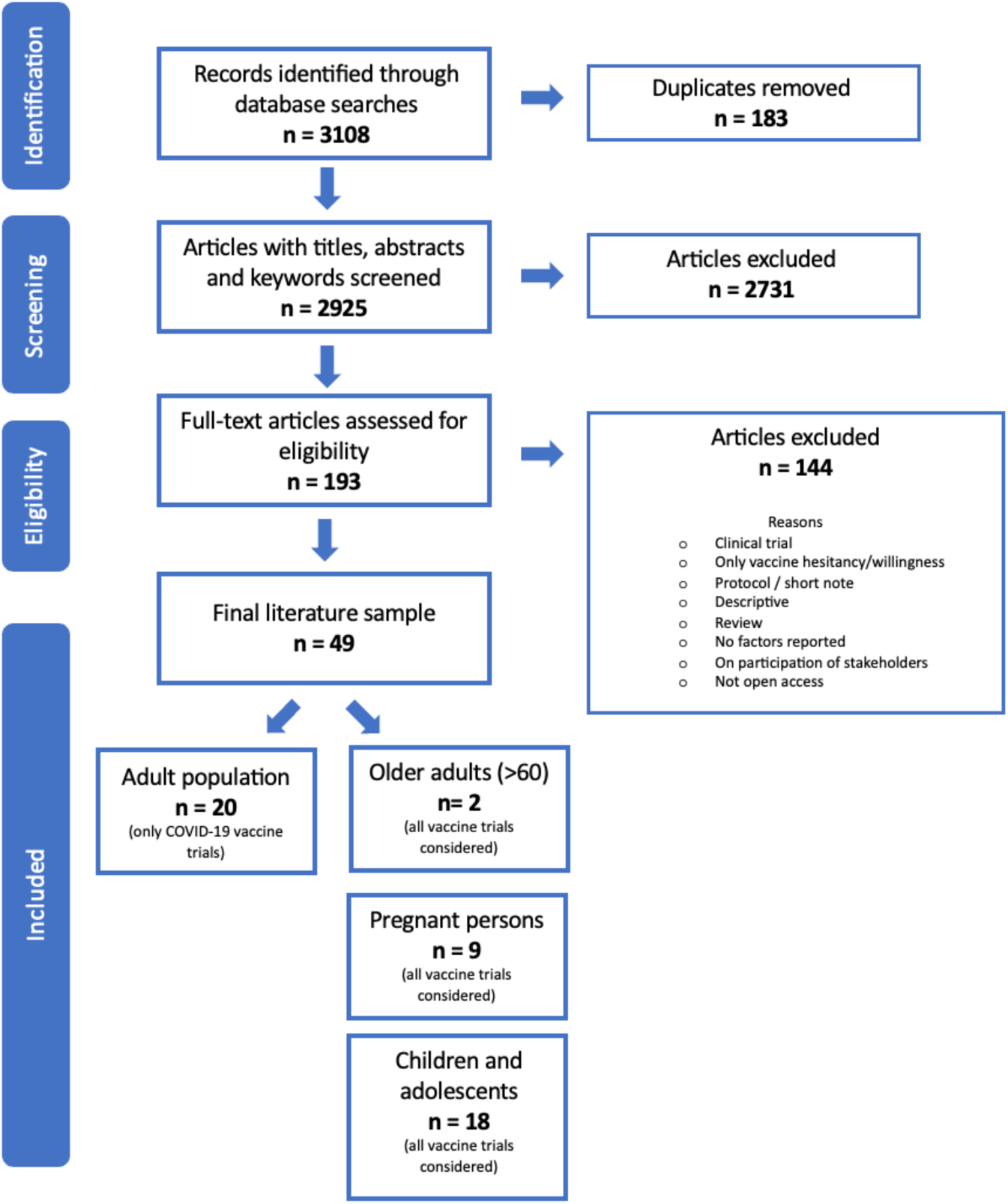
Flowchart identification of articles

**Table 1:** Summary of included articles.

| Authors | Country | Date conducted | Disease of interest | Method | Population | Sample size | Percentage WTP (%) |
| --- | --- | --- | --- | --- | --- | --- | --- |
| General population (only COVID-19 vaccine trials) |  |  |  |  |  |  |  |
| Kitonsa et al. (2021) | Uganda | Sep – Nov 2020 | COVID-19 | Online survey | HCPs | 657 | 70.2 |
| Thompson et al. (2021) | USA | Jun – Dec 2020 | COVID-19 | Online/ telephone survey | Adults | 1835 | 25 |
| Alhassan et al. (2021) | Ghana | 18 Sep – 23 Oct 2020 | COVID-19 | Online survey | Adults | 1556 | 39.8 |
| Felemban et al. (2021) | Saudi Arabia | Sep 2020 | COVID-19 | Online/ telephone survey | Adults | 614 | 40.2 |
| Elshammaa et al. (2022) | Egypt, Saudi Arabia | Oct – Nov 2020 | COVID-19 | Online survey | Adults | 800 | 17.5 |
| Abu-Farha et al. (2020) | Jordan | First half 2020 | COVID-19 | Online survey | Adults | 1287 | 36.1 |
| Echoru et al. (2021) | Uganda | Jul – Sep 2020 | COVID-19 | Online survey | Adults | 1067 | 44.6 |
| Ekezie et al. (2021) | UK | Jul – Aug 2020 | COVID-19 | Focus groups & semi-structured interviews | Different ethnic and vulnerable groups | 70 | / |
| Abdelhafiz et al. (2021) | Egypt, Jordan, Saudi Arabia | 27 Jul – 4 Aug 2020 | COVID-19 | Online survey | Adults | 1576 | 43.9 |
| Sethi et al. (2021) | UK | 4 Sep – 9 Oct 2020 | COVID-19 | Online survey | Adults | 4884 | 41.4 |
| Sun et al. (2021) | China | 20 Mar – 10 Apr 2020 | COVID-19 | Online survey | University students | 1912 | 64.0 |
| Wentzell and Racila (2021) | USA | Sep – Nov 2020 | COVID-19 | Semi-structured phone interviews | Adults<sup>oo</sup> | 31 | / |
| Lusk et al. (2022) | USA | 10 Apr – 30 Jun 2021 | COVID-19 | Online survey | HCPs | 24,769 | 62.2 |
| Gagneux-Brunon et al. (2022) | France | 10-23 May 2021 | COVID-19 | Online survey | Adults | 3,056 | 28.0 |
| Ridde et al. (2021) | Senegal | 24 Dec 2020 – 16 Jan 2021 | COVID-19 | Telephone survey | Adults | 607 | 44.3 |
| Kasozi et al. (2021) | Uganda | 5 Sept – 7 Oct 2020 | COVID-19 | Online survey | HCPs and staff | 260 | 15.8 |
| Jain et al. (2021) | India | 2 Feb – 7 Mar 2021 | COVID-19 | Online survey | Medical students | 1068 | 49.1 |
| Detoc et al. (2020) | France | 26 Mar – 20 Apr 2020 | COVID-19 | Online survey | Adults | 3259 | 47.6 |
| McElfish et al. (2021) | USA | 27 Jul – 22 Nov 2020 | COVID-19 | Online survey | Adults | 120 | 16.7 |
| Lucia et al. (2021) | USA | / | COVID-19 | Online survey | Medical students | 168 | 52.8 |
| Target group: Older adults (>60) |  |  |  |  |  |  |  |
| Raheja et al. (2018) | USA | Nov 2016 – Apr 2017 | / | Self-administered offline/telephone survey | 60+ | 400 | 64.3 |
| Rikin et al. (2017) | USA | 2016 | Influenza | Researcher administered survey | 65+ | 200 | 36 |
| Target group: Pregnant women |  |  |  |  |  |  |  |
| Jaffe et al. (2020) | USA | May – Aug 2017 | Zika | In-depth semi-structured interviews | (recently) Pregnant women | 13 | / |
| McQuaid, Pask, et al. (2016) | UK | Mar – May 2014 | GBS | Interviews and focus groups | Pregnant women, mothers of children with GBS, maternity professionals | 50 | / |
| Goldfarb et al. (2018) | USA | May – Aug 2017 | Zika | Survey | Pregnant and recently postpartum persons | 128 | 77 |
| Karafillakis et al. (2021) | France, Germany, Italy, Spain, UK | Feb – May 2019 | / | Semi-structured interviews and focus groups | Pregnant women | 258 | / |
| Anderson et al. (2021) | UK | 24 Apr – 7 May 2020 | COVID-19 | Semi-structured interviews (telephone/ videoconference) | Pregnant women | 31 | / |
| McQuaid et al. (2018) | UK | Oct 2014 – Jul 2015 | GBS | Survey | Pregnant women & HCPs | 639 | 23.8 |
| Wilcox et al. (2019) | UK | Jul 2017 – Jan 2018 | RSV | Survey | Pregnant women & HCPs | 525 | 28 |
| McQuaid, Jones, et al. (2016) | UK | Sep 2013 | GBS | Survey | Persons of childbearing age | 1013 | 42 |
| Palmer et al. (2016) | Canada | Jun – Jul 2015 | / | Survey | Pregnant women | 112 | 16.3 |
| Target group: Children and adolescents |  |  |  |  |  |  |  |
| Yilmaz and Sahin (2021) | Turkey | 8 – 21 Feb 2021 | COVID-19 | Survey | Parents | 1035 | 4.4 (children)<br>21.0 (parents) |
| Chapagain et al. (2022) | Nepal | Sep – Dec 2020 | / | Survey | Parents | 201 | 67.7 |
| Achieng et al. (2020) | Kenya | Jul 2016 – Aug 2018 | Malaria | Focus groups & interviews | Parents, community members, and study clinicians | 112 | / |
| Buregyeya et al. (2015) | Uganda | Jun – Jul 2008 | Tuberculosis | Focus groups & interviews | Adolescents, parents, and community leaders | / | / |
| Clausen et al. (1954) | USA | 1954 | Polio | Interviews | Parents (mothers) | 175 | / |
| Cunningham et al. (2015) | USA | Jan – Apr 2015 | HPV | Survey | Parents | 256 | 14.5 |
| Hoover et al. (2000) | USA | Sep 1998 | HPV | In-person survey | Female adolescents | 60 | 30 |
| Erves et al. (2017) | USA | Nov 2014 – May 2015 | HPV | Survey | Parents of adolescents | 256 | 47 |
| Farquhar et al. (2006) | Kenya | Dec 2003 – Apr 2004 | HIV | In- person survey | Pregnant women | 805 | 91 |
| Giocos et al. (2008) | South Africa | 2006 | HIV | Survey | Adolescents | 194 | / |
| Goldman et al. (2021) | Canada, USA, Israel, Japan, Spain, Switzerland | Mar – Jun 2020 | COVID-19 | Survey | Parents | 2775 | 18.4 |
| Guerra et al. (2012) | Puerto Rico | 2006 (1)<br>2009 – 2010 (2) | Dengue | Focus groups & interviews | Children & parents | 326 | / |
| Jaspan et al. (2006) | South Africa | Aug 2004 – Mar 2005 | HIV | Survey | Adolescents | 356 | 79 |
| Mahomed et al. (2008) | South Africa | / | Tuberculosis | Survey & focus groups | Adolescents & parents | 270 | / |
| Middelkoop et al. (2008) | South Africa | Nov 2003 – Apr 2004 | HIV | Survey | Adolescents & adults | 200 | 40 |
| Otwombe et al. (2011) | South Africa | Oct 2008 – Mar 2009 | HIV | Survey | Adolescents | 506 | 75 |
| Williamson et al. (2017) | South Africa | Oct 2012 – Jan 2013 (1)<br>Oct 2015 – Mar 2016 (2) | HIV | Interviews | Parents (mothers) | 22 | / |
| Jay et al. (2007) | UK | Jan – Nov 2003 | / | Survey | Parents | 187 | / |

Abbreviations: WTP: Willingness to participate; HIV: Human Immunodeficiency Virus, HPV: Human Papillomavirus, GBS: Group B Streptococcus, RSV: Respiratory Syncytial Virus, HCP: Healthcare Provider.

A total of 3,108 articles were identified through initial database searches. After removing duplicates, 2,924 unique publications remained, of which one member of the research team screened the titles, key words, and abstracts to assess eligibility for inclusion. We excluded review articles, as well as studies that addressed the willingness to participate in non-COVID-19 vaccine trials (unless the study population specifically involved one of our pre-specified target populations). Following this, the full text of 193 articles was reviewed, of which 49 met the inclusion criteria in our scoping review (*Figure 1*). The final set of unique articles comprised 20 articles addressing COVID-19 vaccine trial participation in the general population. The remaining 29 articles provided an evaluation of the willingness to participate in vaccine trials in general (not limited to COVID-19) among our three pre-specified target populations: older adults (n = 2), pregnant women (n = 9) and children and adolescents (n = 18).

The following items were collected for all articles combined (as available in each article): study setting (country), study design, study population, sample size, timing of data collection, and the reported proportion of participants willing to participate in vaccine trials. In addition to these foundational data, we placed a strong emphasis on identifying and analysing the barriers to, and facilitators of participation in COVID-19 vaccine trials, as well as overall vaccine trials, specifically for three target groups: pregnant women, children and adolescents, and older adults. To analyse the factors influencing participation in vaccine trials, we conducted an inductive thematic analysis following the approach described by Braun and Clarke (2006). This involved reviewing the full texts of all included articles, identifying relevant text segments, and coding these inductively to capture key barriers and facilitators. The extracted codes were then grouped into broader themes that reflected recurring patterns across the literature. These themes were organised into distinct categories to highlight the specific factors influencing participation in vaccine trials. This qualitative approach enabled a comprehensive synthesis of findings. This review was aimed to be primarily descriptive in nature.

## Results

### Study characteristics and context

*Table 1* presents an overview of the 49 studies included in this scoping review, detailing key attributes such as the country of origin, timeframe of data collection, study design, target population, sample size, and, where available, the percentage of participants willing to join a vaccine trial. The table is organised into groups: the general population, with a focus exclusively on COVID-19 vaccine trials, and three underrepresented groups: older adults, pregnant women, and children and adolescents.

### Barriers to COVID-19 vaccine trial participation

**Safety concerns** were commonly cited as a major barrier to participation. Multiple studies highlighted apprehensions about the safety of the vaccine candidates and the trials themselves. Participants expressed concerns about exposure to unproven vaccines, potential severe side effects, and a general notion of fear surrounding vaccine trials (Felemban et al., 2021; Kitonsa et al., 2021; Lucia et al., 2021; Ridde et al., 2021). Some also referenced past instances where COVID-19 vaccine trials were paused due to safety concerns, which heightened the sense of risk. Additionally, minor risks, such as experiencing adverse events at the injection site, discouraged some initially willing participants (Ekezie et al., 2021; Kitonsa et al., 2021).

Closely related to safety concerns, **lack of knowledge** also played a significant role in dissuading participation. Participants reported fears of contracting COVID-19 from the vaccine itself (Abdelhafiz et al., 2021; Abu-Farha et al., 2020; Kitonsa et al., 2021), and confusion about trial procedures, often attributable to a general lack of understanding about the nature and outcomes of the trials (Abdelhafiz et al., 2021; Alhassan et al., 2021; Ekezie et al., 2021; Kasozi et al., 2021; Ridde et al., 2021). A qualitative study that specifically addressed concerns of vulnerable groups in the society, such as ethnic minorities and individuals with mental health concerns, showed they were particularly uncomfortable with the idea of visiting hospitals for research, fearing infection or inadequate follow-up care (Ekezie et al., 2021). In some cases, language barriers and insufficient communication also exacerbated fears, leading to greater reluctance (Ekezie et al., 2021).

Another factor was **practical constraints**, which particularly affected healthcare providers and individuals with demanding schedules. Respondents across studies expressed concerns about the time commitment required for trial participation, especially if frequent clinic visits were involved over extended periods (Abu-Farha et al., 2020; Kitonsa et al., 2021; Sun et al., 2021). For many, the logistical challenge of balancing trial participation with professional responsibilities made them less willing to engage.

**Mistrust** emerged as a multifaceted barrier. Participants voiced concerns about the rapid development of COVID-19 vaccines, suspecting that pharmaceutical companies, public health authorities, or medical staff involved in conducting vaccine trials might not have their best interests in mind (Abdelhafiz et al., 2021; Abu-Farha et al., 2020; Alhassan et al., 2021; Ekezie et al., 2021; Gagneux-Brunon et al., 2022; Jain et al., 2021; Kasozi et al., 2021; Ridde et al., 2021; Sun et al., 2021). Mistrust was particularly pronounced in minority groups, with some expressing fears of being exploited or used as “lab rats” (Abdelhafiz et al., 2021; Ekezie et al., 2021; Gagneux-Brunon et al., 2022). For example, in a US study, medical mistrust, particularly influenced by experiences of discrimination, was shown to significantly decrease willingness to participate in vaccine trials among Black participants, who expressed doubts about receiving equitable care compared to other racial groups (Thompson et al., 2021). This mistrust extended to suspicions about the true intentions behind vaccine development, with some believing development was politically or commercially motivated, further deterring participation (Abdelhafiz et al., 2021; Ekezie et al., 2021; Felemban et al., 2021). These findings confirm the importance of the notion of trust as an essential element for the effectiveness of health systems and for the intention to adopt health-promoting behaviours.

In addition to mistrust, **personal, religious, spiritual, cultural, or political beliefs** also shaped participation decisions. One study allowed participants to freely express (they were given an open textbox) reasons for non-participation, with several citing political connotations or religious concerns about vaccine ingredients as their main reason for hesitation (Alhassan et al., 2021). For example, some South Asian respondents were concerned about the presence of animal products in the vaccines, furthermore, this group also acknowledged the impact of cultural views: “*I live in a multicultural society where often cultural views distort scientific facts, so people would not trust science and data even if it shows that there is minimal risk”.* Another example of such a cultural view was found among Gypsy and Roma communities, who expressed fatalistic attitudes towards health, viewing vaccines as unnecessary (Ekezie et al., 2021). These beliefs, intertwined with cultural scepticism towards science and government institutions, contributed to reduced participation.

Finally, **personal stake**, particularly the perceived lack of direct benefits, was mentioned as a reason for not participating. Participants felt that the potential sacrifices required for trial participation, such as avoiding pregnancy or undergoing blood tests, outweighed the personal benefits, especially when there was no guarantee of receiving the vaccine rather than a placebo (Abdelhafiz et al., 2021; Felemban et al., 2021; Kitonsa et al., 2021). This lack of immediate personal gain, combined with uncertainties around vaccine efficacy, led to decreased willingness to participate in trials.

### Facilitators to COVID-19 trial participation

**Altruism, the desire to help others,** was a significant motivator for participation in COVID-19 vaccine trials (Abdelhafiz et al., 2021). Many respondents viewed their potential involvement as a contribution to the fight against the pandemic. For instance, a large segment of the French adult population expressed a sense of duty to participate to help combat COVID-19 (Gagneux-Brunon et al., 2022). Similarly, a study involving young adults in China found that prosocial behaviours were positively correlated with willingness to participate in vaccine trials [19]. Senegalese participants also articulated a desire to “set a good example” as a motivating factor for their participation (Ridde et al., 2021). However, as one study noted, the potential influence of a social desirability bias should be kept in mind (Gagneux-Brunon et al., 2022). At the same time, willingness to support scientific progress, which can be seen as an extension of altruism, was at play. Participants in a qualitative study expressed that their involvement in COVID-19 vaccine trials represented ‘support for the enterprise of science’, many viewed their participation as contributing to what they considered an ‘unprecedented scientific achievement’ [20]. Other studies indicated that the motivation to assist science, alongside a desire to restore normalcy, was among the most significant drivers for participation in vaccine trials (Abdelhafiz et al., 2021; Abu-Farha et al., 2020; Felemban et al., 2021). Qualitative findings also suggested that the aspiration to support scientific endeavours inspired participants to engage in other socially beneficial activities, such as blood donation and health research participation (Wentzell & Racila, 2021).

**Trust in vaccines, healthcare providers, and the government** was another essential facilitator. A lack of trust has been shown to deter individuals from joining vaccine trials, while the presence of trust significantly motivated participation. Research revealed that confidence in vaccines and healthcare providers ranked among the top factors encouraging participation in COVID-19 vaccine trials (Ridde et al., 2021; Sethi et al., 2021). A positive attitude towards vaccination and belief in government capabilities to manage the pandemic were also strongly associated with increased willingness to participate (Ridde et al., 2021). Moreover, actual participants in vaccine trials reported that their belief in science and vaccines was integral to their identity and motivated their involvement (Wentzell & Racila, 2021).

**Incentives**, whether monetary or related to medical care, also served as a motivating factor for participation. Many individuals expressed that the prospect of receiving free medical care (Felemban et al., 2021; Kitonsa et al., 2021) or additional healthcare benefits (Abdelhafiz et al., 2021) played a significant role in their decision to participate in a vaccine trial. The importance of the monetary aspect, including direct financial compensation, was also highlighted as an incentive to join vaccine trials (Abdelhafiz et al., 2021; Kitonsa et al., 2021). However, qualitative interviews with participants of an ongoing COVID-19 vaccine trial revealed that monetary incentives were not a primary motivator for those who took part in the trial (Wentzell & Racila, 2021).

**Calculation**, the **cognitive process** people use to **weigh the perceived risks and benefits,** was a crucial aspect that motivated individuals to consider vaccine trial participation. Deliberate decisions based on the risks associated with COVID-19, coupled with the potential benefits of vaccination, encouraged participation in trials (Detoc et al., 2020; Jain et al., 2021; Ridde et al., 2021; Sun et al., 2021). Personal experiences, such as having had COVID-19 or knowing someone who suffered from severe illness due to the virus, significantly heightened awareness of individual risk (Felemban et al., 2021). Furthermore, concerns about protecting family members from COVID-19 also emerged as a strong motivator for involvement in vaccine trials (Abdelhafiz et al., 2021; Abu-Farha et al., 2020; Kitonsa et al., 2021).

**Targeted (or culturally adapted) information** on the trial process (including transparency at all stages, vaccine content, and reflecting cultural appropriateness) emerged as a critical facilitator, especially for ethnic minorities and vulnerable populations (Echoru et al., 2021; Ekezie et al., 2021; Lusk et al., 2022). For example, Muslim participants highlighted the need for culturally relevant information regarding vaccine ingredients to ensure they were halal and free from alcohol and animal products. Additionally, these groups required reassurances regarding hospital safety and the neutral settings used for research to alleviate fears of infection (Ekezie et al., 2021). Clear evidence of diverse ethnic participation in trials was particularly vital for African and African Caribbean communities, as it helped mitigate feelings of isolation in their involvement (Ekezie et al., 2021). In the same study, other communities, like the Gypsy and Roma groups, called for more culturally sensitive health communication (Ekezie et al., 2021).

### Barriers and facilitators in pre-specified target groups

#### Older adults: barriers and facilitators for vaccine trial participation

Only two articles dealt with barriers and facilitators for vaccine trial participation in the older adult population. Barriers to participation included not sharing or **not being informed about the results of the trial** (Raheja et al., 2018), **distance to the research facility** (Raheja et al., 2018), and, maybe surprisingly, **monetary incentives** (Rikin et al., 2017). While monetary incentives were mentioned before as a facilitator, they acted as a barrier for older adults, who saw them as pressure tactics rather than a sincere incentive to participate. On the contrary, respondents were more likely to participate in a vaccine trial if they had already **participated in a trial in the past** or if they had more **prior knowledge about vaccine trials** (Raheja et al., 2018; Rikin et al., 2017). Also, respondents with a **better health status** were more likely to participate in a vaccine trial (Raheja et al., 2018).

#### Pregnant women: barriers and facilitators for vaccine trial participation

For pregnant women, **safety concerns** were overwhelmingly the most significant barrier for participation. Pregnant women expressed apprehension regarding the unknown risks associated with experimental vaccines, particularly their effects on foetal health. A study investigating willingness to participate in Zika vaccine trials revealed that, among persons who were unsure about vaccine safety for their baby or who believed the vaccine to be “likely or definitely not safe” for the baby, a majority (ranging from 56%-97% over different scenarios) declined trial participation, highlighting safety of the baby being the top priority (Goldfarb et al., 2018). Qualitative research identified concerns about experimental products, potential (long-term) side effects, and risks to babies as common reasons for refusal: “*It’s a study where you try to find out the effects of a certain product on pregnant women and their babies. You don’t know the risks yet, you will see the effects during the trial and I am not willing to put the health of my baby in danger.*” (Karafillakis et al., 2021). These worries were echoed in other studies, with particular emphasis on miscarriage risk (McQuaid et al., 2018), the feeling of no or little individual benefits (Karafillakis et al., 2021; Palmer et al., 2016), and the perception that contracting COVID-19 posed less danger than vaccination (Anderson et al., 2021). Moreover, maternity professionals were hesitant to recommend trial participation due to safety concerns and potential legal issues (McQuaid et al., 2018; McQuaid, Pask, et al., 2016). Additionally, a study showed that willingness to participate varied by vaccine type, with higher acceptance for inactivated virus and nucleic acid-based vaccines and lower willingness for live-attenuated vaccines (Goldfarb et al., 2018).

Additionally, there was a pronounced **need for more information** regarding vaccine trials (Anderson et al., 2021; Karafillakis et al., 2021; McQuaid, Pask, et al., 2016). Studies reported low awareness levels about participation options, and persons who showed willingness to participate often sought additional details about the trial processes and safety measures for their specific situation (McQuaid, Pask, et al., 2016).

Lastly, also **mistrust** and even disgust regarding the pharmaceutical industry, were mentioned by respondents of several studies as reasons why they would not participate (Anderson et al., 2021; Karafillakis et al., 2021; Palmer et al., 2016). Many persons expressed scepticism about the motives behind vaccine development, perceiving it as profit-driven rather than genuinely aimed at public health. In one study a respondent mentioned: ‘‘*I think there is a lot of vested interests behind all this, the pharmaceutical business, an economic benefit, and I don’t want to take part in any of that. In fact, it’s disgusting*” (Karafillakis et al., 2021).

On the other hand, several facilitators could motivate pregnant women to participate in vaccine trials. The **assurance of safety** emerged as a crucial factor. Participants indicated they would only consider involvement in later trial phases after safety data were available from prior studies (Anderson et al., 2021; Karafillakis et al., 2021). In another study, pregnant women indicated that they would like to see evidence of safety from others who had fallen pregnant during their participation in a vaccine trial (Goldfarb et al., 2018). The significance of the notion of safety is further illustrated by the observation that the majority of persons who believed the vaccine was definitely or likely safe for their baby, accepted trial participation (Goldfarb et al., 2018). Additionally, some pregnant women were motivated by the possibility of being in a control group, allowing them to contribute to research without perceived risks to their babies (McQuaid, Pask, et al., 2016). Once again, this highlights the importance of informing individuals about vaccine safety. While the concept of a randomised controlled trial model may encourage participation (McQuaid, Pask, et al., 2016), the opposite effect can be observed in pregnant women who were already willing to participate in a vaccine trial.; one study showed they became less inclined to participate as they sought a guarantee of receiving the vaccine (McQuaid et al., 2018).

Pregnant women showed a greater likelihood of participating in vaccine trials when their **risk perception** of the disease was higher. Among participants who were willing to participate in a vaccine study, 65% cited their desire to protect themselves, while almost all (98%) cited a desire to protect their babies as motivating factors (Goldfarb et al., 2018; McQuaid, Pask, et al., 2016). Previous experiences with disease heightened willingness to participate, as one respondent expressed a readiness to volunteer based on her child’s previous illness (McQuaid, Pask, et al., 2016; Wilcox et al., 2019). This risk-benefit analysis was notably significant in trials for life-threatening diseases. In a COVID-19 vaccine trial study, several participants indicated a willingness to volunteer if they perceived a higher risk of the virus to pregnant women (Anderson et al., 2021). Engaging parents affected by the disease or using patient advocates to discuss risks related to the disease with pregnant women (or maternity professionals) were suggested as strategies to enhance participation (McQuaid, Pask, et al., 2016).

**Incentives** linked to medical care significantly motivated pregnant women to participate in vaccine trials, including additional scans, antenatal appointments, and extended healthcare for both parent and child (McQuaid et al., 2018; McQuaid, Pask, et al., 2016; Palmer et al., 2016). However, based on the literature we reviewed, financial compensation was not identified as a significant motivator for participation among pregnant women, as only a small percentage of participants found monetary payments to be compelling (Karafillakis et al., 2021; Palmer et al., 2016).

Receiving comprehensive information, thereby increasing **vaccine trial literacy**, about the trial procedures and safety was crucial for encouraging participation among pregnant women. Recommendations from healthcare providers (Goldfarb et al., 2018; Karafillakis et al., 2021), particularly midwives, were also influential, with many preferring invitations from their midwives (McQuaid et al., 2018; McQuaid, Pask, et al., 2016). Furthermore, enthusiasm and support from the midwife about the trial would motivate the pregnant woman to participate (McQuaid, Pask, et al., 2016).

While some participants expressed a willingness to participate for **altruistic** reasons and to advance scientific knowledge, this was not the dominant motivator for most (Anderson et al., 2021; Karafillakis et al., 2021; McQuaid, Pask, et al., 2016). A few participants from various countries acknowledged the importance of trials in improving safety for future pregnancies (Karafillakis et al., 2021). In a different study someone noted her readiness to participate in trials, after considering her own health and age, if it could help other pregnant women in the future (Anderson et al., 2021).

#### Children and adolescents: barriers and facilitators for vaccine trial participation

The willingness to participate in vaccine trials was questioned through parental attitudes in most studies. This approach is logical, as parents often have the final say in whether their children and adolescents take part in vaccine trials. Beyond influencing this decision, they are also responsible for providing informed consent, just as they are in many cases the primary decision-makers for actual vaccination agreement as well. However, some studies also questioned adolescents’ way of reasoning on the topic, leading to a diverse set of factors that can either hinder or encourage participation.

For children and adolescents as a target group, also, vaccine trial **safety concerns** appear to be an often-mentioned barrier to (let them) participate in vaccine trials (Abu-Farha et al., 2020; Buregyeya et al., 2015; Chapagain et al., 2022; Farquhar et al., 2006; Guerra et al., 2012; Mahomed et al., 2008; Williamson et al., 2017). Parents often worry about potential side effects, such as their children becoming sick, developing allergic reactions, or some even mentioned the fright of their child dying. In an HIV vaccine trial, 80% of mothers cited fear of side effects as the primary reason for not participating (Farquhar et al., 2006). Similar concerns were raised in tuberculosis (TB) and dengue vaccine trials, where parents feared their children would contract the diseases from the vaccine itself (Buregyeya et al., 2015; Guerra et al., 2012; Mahomed et al., 2008).

**Mistrust** of vaccines, vaccine trials, and researchers is another important barrier. Parents frequently express distrust of new vaccines and the procedures used in clinical trials, with some feeling that trials exploit vulnerable communities. In a dengue vaccine trial, parents said they would not trust what was being injected into their children (Guerra et al., 2012). Similarly, participants in TB vaccine trials questioned why researchers didn’t test the vaccine on themselves first, with some fearing they were being used as “guinea pigs” (Buregyeya et al., 2015). This mistrust often stems from experiences with medical discrimination and racial biases, with concerns that vaccines might harm fertility or reduce lifespans. Additionally, some parents perceive incentives like monetary compensation or improved medical care as coercive, rather than genuine facilitators for participation (Buregyeya et al., 2015; Guerra et al., 2012).

A **lack of information** also hinders participation. Participants often describe feeling uninformed about the vaccine, trial process, and results of previous studies. This information gap, coupled with lower levels of formal education, can make it challenging for parents to understand the purpose and safety of the research (Buregyeya et al., 2015; Guerra et al., 2012). Continuous and clear communication throughout the trial can help overcome these barriers by building trust and fostering understanding.

Both parents and adolescents express a desire to contribute to science and help the community, highlighting **altruism** as a facilitator. Mothers, for example, have cited the wider community benefits as a reason to enrol their child in trials, hoping to reduce diseases and help other children (Williamson et al., 2017). Adolescents, too, express altruistic motivations. One study questioning adolescents between 11- and 19-years old reports altruistic reasoning as the main facilitator leading them towards willingness to participate in vaccine trials (Jaspan et al., 2006). Altruism was also highlighted in an HPV vaccine trial, where nearly 47% of respondents said the potential benefit for women strongly influenced their decision to participate (Hoover et al., 2000). Parental willingness to include their child in a vaccine trial is complex, but altruistic motives are a factor strongly associated with their choice to participate (Goldman et al., 2021).

**Trust** in medical researchers is another important facilitator. A good relationship between participants and researchers can foster trust, increasing willingness to participate in trials. For example, in an HPV vaccine trial, higher trust in medical researchers was associated with greater parental willingness to enrol their children (Erves et al., 2017). Trust can be built through good communication, both verbal and nonverbal, as well as through interactions with medical experts who provide guidance throughout the trial process (Chapagain et al., 2022; Goldman et al., 2021; Jay et al., 2007). Maintaining this trust is key to retaining participants in vaccine trials.

Multiple studies highlighted the importance of **vaccine trial literacy**, referring to the ability to understand vaccine trial information. In one study, parents and adolescents with a higher level of understanding of the vaccine, the disease, and the trial procedures were more willing to participate [50]. For example, in a study on TB vaccines, participants who received clear explanations of the trial’s risks were more likely to join future vaccine trial studies (Mahomed et al., 2008). Similarly, better knowledge of HIV and a hypothetical HIV-vaccine was linked to higher retention in future trials, possibly due to educational programs that improved participants’ understanding (Middelkoop et al., 2008). Addressing the information gap by providing easy-to-understand, accessible information can significantly enhance participation rates (Achieng et al., 2020; Clausen et al., 1954; Erves et al., 2017).

**Parental willingness to participate in trials** also plays a role in increasing participation among children and adolescents. Mothers who expressed positive attitudes towards joining trials themselves were also more likely to allow their children to participate (Farquhar et al., 2006). Another dynamic was observed in COVID-19 vaccine trials, where parental willingness to receive the vaccine was the most critical determinant of their children’s involvement (Goldman et al., 2021; Yilmaz & Sahin, 2021). Identifying those parents with positive attitudes towards both vaccines and vaccine trials can therefore be a promising strategy to increase the level of recruitment for children and adolescents.

Finally, **incentives**, both medical and monetary, motivated participation. The prevention of the disease was a key motivator, the more since it included free consultation and medication (Achieng et al., 2020). But also, the discovery of other unknown health conditions during the vaccine trials and a treatment for these, contributed to parental willingness (Guerra et al., 2012), particularly for those who may not have regular access to healthcare. Monetary incentives, such as compensation for transportation and additional payments at the end of the trial, are also influential, with suggestions ranging from allowances to mosquito nets, food, clothing, and school fees (Achieng et al., 2020; Guerra et al., 2012; Hoover et al., 2000). However, it is important to note that while incentives can encourage participation, they may also be perceived as coercive by some parents, potentially reducing their willingness to enrol their children (Buregyeya et al., 2015; Guerra et al., 2012).

## Discussion

This scoping review provides new insights into the complex and often underexplored factors that influence participation in vaccine trials, with a dual focus: participation in COVID-19 vaccine trials among the general population and overall vaccine trial participation among three underrepresented groups: pregnant women, children and adolescents, and older adults (>60). By identifying key barriers and facilitators, this review contributes to a broader understanding of how participation in vaccine trials can be improved, ultimately supporting more inclusive and effective vaccine development strategies.

The review followed a primarily descriptive and thematic approach, with relevant data extracted from the selected literature and analysed using an inductive thematic analysis, as described in the methods section. Given the limited availability of studies specifically addressing these target groups in the context of COVID-19 vaccine trials, broader vaccine trial literature was also considered. This approach allowed for a more comprehensive understanding of participation dynamics across different populations, ensuring that insights gained could be applied to both pandemic preparedness and routine vaccine development. In doing so, this review aimed to support the expansion of vaccine trial volunteer registries and provide guidance for future recruitment efforts.

The barriers and facilitators for participation in (COVID-19) vaccine trials highlight the complex interplay of factors influencing the decision to participate. From safety concerns to altruistic motivations, the factors influencing trial participation vary widely, but some recurring themes stand out. *Table 2* provides an overview of the key barriers and facilitators identified across the literature, serving as a visual summary of the themes discussed in this review.

**Table 2:** Overview identified barriers and facilitators.

| Barriers | Description |
| --- | --- |
| Safety concerns | ○ Safety concerns vaccine candidates<br>○ Side effects<br>○ Perceived rapid development |
| Lack of knowledge | ○ Fear of contracting disease during vaccine trial<br>○ Confusion trial procedures<br>○ Fear of inadequate follow-up |
| Practical constraints | ○ Time constraints<br>○ Logistical challenges with job<br>○ Distance research facility |
| Mistrust | ○ Mistrust in pharmaceutical companies, public health authorities, medical staff conducting vaccine trials<br>○ Medical discrimination<br>○ Development vaccines politically or commercially motivated |
| Personal, religious, spiritual, cultural, or political beliefs | ○ Concerns vaccine ingredients<br>○ Cultural negative view towards vaccine trials<br>○ Scepticism towards science and government institutions<br>○ Immune system > vaccine, trial unnecessary |
| Personal stake | ○ Lack of personal gain<br>○ No guarantee of receiving vaccine in trial<br>○ Sacrifices: avoiding pregnancy, undergoing blood tests... |
| Lack of information | ○ Low awareness of options<br>○ Search for additional information<br>○ Need information trial process<br>○ Not receiving results of trial |
| Monetary incentives | ○ Perceived pressure tactic<br>○ Raises suspicion |
| Facilitators | Description |
| Altruism | ○ Desire to help others<br>○ Set a good example<br>○ Fight against pandemic<br>○ Support scientific progress<br>○ Restore normalcy |
| Trust | ○ Trust in vaccines<br>○ Trust in healthcare providers,<br>○ Trust in government<br>○ Trust in safety of trials |
| Incentives | ○ Financial compensation<br>○ Receiving free medical care<br>○ Medical follow-up parent and child<br>○ Help in treatment unknown health conditions<br>○ Others: food, clothing, school fees... |
| Calculation | ○ Deliberate decisions<br>○ Risk-benefit balance<br>○ Personal experiences<br>○ Prior participation |
| Targeted information | ○ Culturally adapted information<br>○ Transparency<br>○ Targeted to different groups |
| Health status | ○ Better health status |
| Assurance of safety | ○ Availability safety data<br>○ First-hand experiencers<br>○ Perceived vaccine safety drives perceived trial safety<br>○ RCTs can motivate but also discourage participation |
| Vaccine trial literacy | ○ Understanding trial procedures<br>○ Understanding safety in trials<br>○ Prior knowledge<br>○ Comprehensive information from HCPs |
| Risk perception disease | ○ Higher risk perception<br>○ Desire to protect oneself and others<br>○ Previous experience disease |
| Parental willingness | ○ Parents positive trial attitudes<br>○ Willingness to receive vaccine |

One of the most consistently reported barriers to participation, across all groups, are the concerns about vaccine trial safety. This concern was particularly pronounced among pregnant women and parents deciding for their children. Across studies, pregnant women often expressed fear about unknown long-term side effects and potential risks to their unborn child, leading to heightened caution and reluctance. These concerns were amplified by the perceived novelty and speed of development of vaccines.

Mistrust also emerged as a central barrier across groups, which was manifested in multiple forms: mistrust in pharmaceutical companies, public health authorities, medical professionals, and the trial process itself. As highlighted throughout this review, continuous and transparent communication before and during vaccine trials can be essential for overcoming barriers, fostering trust, and ensuring informed participation. Mistrust was particularly evident in marginalised communities, who expressed scepticism about the intentions behind vaccine trials. This mistrust is fuelled by historical deprivation in medical research, as seen in studies where minority participants voiced concerns about being used as “guinea pigs” or targeted for experimentation. Among pregnant women, this mistrust was particularly directed at vaccine manufacturers, who were often perceived as profit-driven rather than genuinely focused on public health. In this context, overcoming mistrust requires more than providing information; it also involves addressing deep inequities in healthcare systems.

On the other hand, several facilitators encouraged participation in vaccine trials, with altruism and trust emerging as consistently reported motivators. Altruistic motivations, such as supporting scientific progress and setting a good example stood out as a powerful driver across all groups. In studies involving adults and adolescents, participants expressed the feeling of wanting to contribute to the greater good by helping to end the pandemic. Trust in vaccines, healthcare providers, and the scientific process emerged as another key facilitator. Participants with high levels of trust were more likely to engage in trials, believing that they were in good hands. In the context of paediatric vaccine trials, targeting parents with favourable views on both vaccines and vaccine research could be an effective approach to improve recruitment of children and adolescents. Overall, ensuring continued trust is vital to maintaining participant involvement in vaccine trials.

Incentives, both monetary and related to healthcare benefits, also played a role in motivating participation. However, qualitative findings suggest that while financial compensation may be appealing, intrinsic motivations rooted in altruism and community support often take precedence.

Furthermore, a recurring facilitator is the balancing of risks and benefits. For many participants, especially those with pre-existing health conditions or experiences with the disease, the potential protective benefits of vaccination outweighed the perceived risks of trial participation. Additionally, the desire to protect family members from a disease also emerged as a strong motivator for involvement in vaccine trials. This underlines the need to emphasise not only the individual health benefits of vaccination but also the broader role of vaccine trials in protecting the community against the virus and its associated disease.

As discussed earlier, safety (concerns) and (mis)trust are significant factors in this review, but addressing the information gap could go a long way in mitigating the challenges that come with these factors. Providing easy-to-understand, accessible information about the vaccine trial process, expected outcomes, and safety measures could significantly enhance participation rates. Ensuring that this information is available in a variety of formats, including those accessible to individuals with different levels of health literacy, is essential for reaching a broad population.

## Conclusion

This review identified several recurring barriers and facilitators to vaccine trial participation. With our findings we aim to inform more effective recruitment strategies. A comprehensive understanding of these factors is essential for designing interventions that enhance trial participation, ultimately accelerating vaccine development and contributing to more equitable public health outcomes.

### Limitations

While search queries were carefully delineated, the exclusive use of PUBMED for literature collection may be seen as a limitation of this review, as this approach could have introduced bias due to the scope of the database. Additionally, older adults were underrepresented in the literature, with only two articles specifically addressing this group. Nonetheless, the searches conducted were extensive and articles were thoroughly screened for eligibility.

Another limitation involved the lack of direct input from children (and limited input of adolescents) in the studies reviewed. Decisions regarding their participation in vaccine trials typically rest with their parents or legal guardians, which may overlook the unique concerns and motivations of children and adolescents. Although parents make the final decision, the willingness and opinions of children and adolescents can significantly influence whether they are permitted to participate. Future research should aim to incorporate their voices, allowing for a more comprehensive understanding of factors affecting their willingness to engage in vaccine trials, that may thus far have gone unrecognised. Ultimately leading to more effective strategies for increasing participation rates among this demographic.

It is important to note that this review does not provide quantitative assessments of the frequency or prevalence of the identified barriers and facilitators. While the thematic analysis offers valuable insights, it remains descriptive in nature. Further research is needed to explore the relationships between these factors and to determine which have the most significant impact on an individual’s willingness to participate in vaccine trials.

### Implications and future research

Addressing factors impacting vaccine trial participation requires a multifaceted approach, with clear, transparent communication being essential. Safety concerns and mistrust (particularly within underrepresented communities) must be alleviated through targeted, accessible messaging that resonates with these populations. Future research should focus on developing and testing communication strategies that directly address these concerns, fostering long-term trust in vaccine trials. Institutions involved in vaccine trials must prioritise proactive and consistent communication to build and maintain trust, ensuring that participants feel informed, valued, and supported throughout the process. In parallel, addressing the historical and ongoing inequities in healthcare is crucial. It is not enough to provide information; efforts must also be made to directly tackle the deeper issues of mistrust that have emerged from past injustices.

Alongside this, leveraging altruism, by framing vaccine trials as opportunities for individuals to contribute meaningfully to public health, can significantly boost engagement. Future research should investigate how altruistic motivations vary across different cultural and demographic contexts to refine recruitment strategies that appeal to those motivated by societal well-being. Healthcare providers and trial organisers can leverage these motivations to encourage broader participation in vaccine trials.

Understanding the balance between financial incentives and other motivators is crucial for participant retention in vaccine trials. While financial compensation can encourage participation, it can also have contradictory effects, acting as both a facilitator and a barrier. Some participants may view incentives positively, while others may see them as exploitative or diminishing the altruistic value of participation. Social desirability bias may also affect how participants report their motivations, potentially skewing results. Future research should explore the long-term impact of financial incentives on participation and trust, and trial organisers should carefully consider how to combine monetary incentives with messages emphasising the broader societal impact of participation.

Finally, future research should explore how individuals weigh the risks and benefits associated with vaccine trials (calculation), particularly among high-risk populations such as pregnant women and older adults. Understanding how these groups calculate the potential risks and benefits will help improve risk communication strategies. In practice, trial organisers should ensure that recruitment materials provide clear and accessible risk-benefit assessments, tailored to the specific concerns of each population. This will help ensure that participants are well-informed and confident in their decision to participate, thereby improving trial participation rates.

These insights and strategies will be instrumental in enhancing the inclusivity and effectiveness of vaccine trials moving forward, ensuring that the trials are accessible, transparent, and appealing to a wide range of participants. This review contributes to the broader field of public health by offering actionable guidance that can support researchers, public health practitioners, and vaccine trial organisers in developing strategies to increase vaccine trial engagement. These efforts will enhance pandemic preparedness by improving vaccine trial participation, facilitating faster testing, and ensuring more timely responses during future pandemics. In turn, this will strengthen global initiatives to combat vaccine-preventable infectious diseases and contribute to more equitable public health outcomes.

## Supporting information

List of searches

## Data Availability

All data produced in the present study are available upon reasonable request to the authors

## Acknowledgments

This work was supported by the VACCELERATE project, which is funded by the European Commission’s activities for future pandemic preparedness under the European Union’s Horizon 2020 research and innovation programme (grant agreement No. 101037867).

## Statement on AI Use

The authors used AI tools (ChatGPT by OpenAI) to support language refinement, structural clarity, and phrasing alternatives during manuscript preparation. AI was also used to explore options for improving the logical flow and coherence of the text. All content decisions, critical interpretation, and final wording were made and approved by the authors. No AI tools were used for data extraction, analysis, or scientific interpretation.

