## Supplementary material for "Building Up Facilitators, Breaking Down Barriers: A Scoping Review Mapping Factors that Impact Participation in Vaccine Trials": List of searches

Log

**Question: Willingness to participate in a Covid-19 trial**

**Search**: COVID*[tiab] AND willingness*[tiab] AND trial*[tiab]

**Search date**: April 11^th^, 2022

**Database**: Pubmed

**Filters used**: Year: 2020-2022; Article type: Books and Documents, Clinical Trial, Meta-Analysis, Observational Study, Randomized Controlled Trial, *Classical Article, Lecture, Letter, Multicenter Study, Congress, Evaluation Study (the ones in italics did not add additional records)*
**Number of records**: 11

**Question: Covid-19 hesitancy and trial participation**

**Search**: COVID*[tiab] AND hesitancy*[tiab] AND trial*[tiab]

**Search date**: April 11^th^, 2022

**Database**: Pubmed

**Filters used**: Year: 2020-2022; Article type: Books and Documents, Clinical Trial, Meta-Analysis, Observational Study, Randomized Controlled Trial, *Classical Article, Lecture, Letter, Multicenter Study, Congress, Evaluation Study (the ones in italics did not add additional records)*
**Number of records**: 10

**Question: Covid-19 hesitancy and trial participation (less filters)**

**Search**: COVID*[tiab] AND hesitancy*[tiab] AND trial*[tiab]

**Search date**: April 11^th^, 2022

**Database**: Pubmed

**Filters used**: Year: 2020-2022
**Number of records**: 131

**Question: Willingness to participate in a covid-19 trial?**

**Search**: COVID*[tiab] AND (particip*[tiab] OR willing*[tiab]) AND trial*[tiab]

**Search date**: April 14^th^, 2022

**Database**: Pubmed

**Filters used**: Year: 2020-2022, Article type: Case Reports, Classical Article, Evaluation Study, Interview, Observational Study, Letter, Lecture
**Number of records**: 2633

**Question: Willingness of children to participate in a covid-19 trial?**

**Search**: COVID*[tiab] AND particip*[tiab] AND (willing*[tiab] OR attitu*[tiab] OR barrier*[tiab] OR facilitat*[tiab]) AND trial*[tiab] AND (child*[tiab] OR pediat*[tiab]) AND (child[MeSH] OR pediatric[MeSH])

**Search date**: April 21^st^, 2022

**Database**: Pubmed

**Filters used**: Year: 2020-2022; Article type: Case Reports, Classical Article, Evaluation Study, Interview, Observational Study, Letter, Lecture (Same articles without filters)
**Number of records**: 20

**Question: Willingness of pregnant women to participate in a covid-19 trial?**

**Search**: COVID*[tiab] AND particip*[tiab] AND (willing*[tiab] OR attitu*[tiab] OR barrier*[tiab] OR facilitat*[tiab]) AND vaccin*[tiab] AND (pregn*[tiab] OR lactat*[tiab] OR perinat*[tiab]) AND trial

**Search date**: April 21^st^, 2022

**Database**: Pubmed

**Filters used**: Year: 2020-2022; Article type: Case Reports, Classical Article, Evaluation Study, Interview, Observational Study, Letter, Lecture (same amount without filters)
**Number of records**: 12

**Question: Willingness of elderly or minorities or adolescent populations to participate in a vaccine trial?**

**Search**: participat*[tiab] AND (willing*[tiab] OR attitu*[tiab]OR barrier*[tiab] OR facilitat*[tiab]) AND vaccin*[tiab] AND trial[tiab] AND ((old[tiab] OR old[MeSH] OR elder*[tiab] OR elder*[MeSH]) OR (minor[tiab] OR minor*[MeSH]) OR (adolesc*[tiab] OR adolesc *[MeSH]))

**Search date**: Jun 20^th^, 2022

**Database**: Pubmed

**Filters used**: /

**Number of records**: 57

**Question: Willingness of at-risk populations to participate in a covid-19 trial?**

**Search**: COVID*[tiab] AND particip[tiab] AND (willing*[tiab] OR attitu*[tiab] OR barrier*[tiab] OR facilitat*[tiab]) AND vaccin*[tiab] AND (risk*[tiab] OR comorb*[tiab] OR cancer[tiab] OR underserv*[tiab] OR underrepr*[tiab])

**OR search**: COVID*[tiab] AND particip[tiab] AND (willing*[tiab] OR attitu*[tiab] OR barrier*[tiab] OR facilitat*[tiab]) AND vaccin*[tiab] AND ((risk*[tiab] OR comorb*[tiab] OR cancer[tiab] OR underserv*[tiab] OR underrepr*[tiab]) OR (risk*[MeSH] OR comorb*[MeSH] OR cancer[MeSH] OR underserv*[MeSH] OR underrepr*[MeSH]))

**Search date**: April 21^st^, 2022

**Database**: Pubmed

**Filters used**: /

**Number of records**: 0

**Question: Willingness of at-risk populations to participate in a vaccine trial?**

**Search**: participat[tiab] AND (willing*[tiab] OR attitu*[tiab] OR barrier*[tiab] OR facilitat*[tiab]) AND vaccin*[tiab] AND trial[tiab] AND ((risk*[tiab] OR risk*[MeSH]) OR (comorb*[tiab] OR comorb*[MeSH]))

**Search date**: Jun 20^th^, 2022

**Database**: Pubmed

**Filters used**: /

**Number of records**: 118

**Question: Willingness of pregnant women to participate in a vaccine trial?**

**Search**: particip*[tiab] AND (willing*[tiab] OR attitu*[tiab] OR barrier*[tiab] OR facilitat*[tiab]) AND vaccin*[tiab] AND (pregn*[tiab] OR lactat*[tiab] OR perinat*[tiab]) AND trial

**Search date**: Jun 20^th^, 2022

**Database**: Pubmed

**Filters used**: /

**Number of records**: 56

**Question: Willingness of children to participate in a vaccine trial?**

**Search**: particip*[tiab] AND (willing*[tiab] OR attitu*[tiab] OR barrier*[tiab] OR facilitat*[tiab]) AND vaccin*[tiab] AND trial*[tiab] AND (child*[tiab] OR pediat*[tiab]) AND (child[MeSH] OR pediatric[MeSH])

**Search date**: Jun 20^th^, 2022

**Database**: Pubmed

**Filters used**: /

**Number of records**: 60

**Total number of searches: 3108** (see flowchart for further identification process of articles)

**Question: COVID-19 hesitancy for at-risk/ underserved populations?**

**Search**: COVID*[tiab] AND (hesitan*[tiab] OR confid*[tiab] OR accept*[tiab]) AND vacc*[tiab] AND (‘at-risk’*[tiab] OR comorb*[tiab] OR cancer[tiab] OR underserv*[tiab] OR underrepr*[tiab] OR old[MeSH])

**Search date**: April 21^st^, 2022

**Database**: Pubmed

**Number of records**:

**Filters used**:

**Saved as**: “Search10.xlsx”

**Eligible based on title**:

**Number of duplicates from previous searches**:

**Eligible based on abstracts:**

COVID*[tiab] AND (hesitan*[tiab] OR confid*[tiab] OR accept*[tiab]) AND vacc*[tiab] AND (‘at-risk’*[tiab] OR comorb*[tiab] OR cancer[tiab] OR underserv*[tiab] OR underrepr*[tiab] OR old*[tiab] OR HIV[tiab] OR elder*[tiab] OR pregnan*[tiab] OR lactat*[tiab] OR child*[tiab] OR pediatric*[tiab]) AND (Europ*[tiab] OR Austria*[tiab] OR Belgi*[tiab] OR Bulgaria*[tiab] OR Croatia*[tiab] OR Cypr*[tiab] OR Czech*[tiab] OR Denmark[tiab] OR Danish[tiab] OR Estonia*[tiab] OR Fin*[tiab] OR France[tiab] OR French[tiab] OR German*[tiab] OR Gree*[tiab] OR Hungar*[tiab] OR Ireland[tiab] OR Irish[tiab] OR Ital*[tiab] OR Latvia*[tiab] OR Lithuania*[tiab] OR Luxemb*[tiab] OR Malt*[tiab] OR Netherlands[tiab] OR Dutch[tiab] OR Poland[tiab] OR Polish[tiab] OR Portug*[tiab] OR Romania*[tiab] OR Slovakia*[tiab] OR Slovenia*[tiab] OR Spain[tiab] OR Spanish[tiab] OR Sweden[tiab] OR Swedi*[tiab])

**Question: Covid-19 hesitancy in EU countries**

**Search query:** COVID*[tiab] AND (hesitan*[tiab] OR confid*[tiab] OR accept*[tiab]) AND vacc*[tiab] AND (Europ*[tiab] OR Austria*[tiab] OR Belgi*[tiab] OR Bulgaria*[tiab] OR Croatia*[tiab] OR Cypr*[tiab] OR Czech*[tiab] OR Denmark[tiab] OR Danish[tiab] OR Estonia*[tiab] OR Fin*[tiab] OR France[tiab] OR French[tiab] OR German*[tiab] OR Gree*[tiab] OR Hungar*[tiab] OR Ireland[tiab] OR Irish[tiab] OR Ital*[tiab] OR Latvia*[tiab] OR Lithuania*[tiab] OR Luxemb*[tiab] OR Malt*[tiab] OR Netherlands[tiab] OR Dutch[tiab] OR Poland[tiab] OR Polish[tiab] OR Portug*[tiab] OR Romania*[tiab] OR Slovakia*[tiab] OR Slovenia*[tiab] OR Spain[tiab] OR Spanish[tiab] OR Sweden[tiab] OR Swedi*[tiab])

**Question: Covid-19 hesitancy in EU countries**

**Search query:** COVID*[tiab] AND (hesitan*[tiab] OR confid*[tiab] OR accept*[tiab] OR **percept***[tiab]) AND vacc*[tiab] AND (Europ*[tiab] OR Austria*[tiab] OR Belgi*[tiab] OR Bulgaria*[tiab] OR Croatia*[tiab] OR Cypr*[tiab] OR Czech*[tiab] OR Denmark[tiab] OR Danish[tiab] OR Estonia*[tiab] OR Fin*[tiab] OR France[tiab] OR French[tiab] OR German*[tiab] OR Gree*[tiab] OR Hungar*[tiab] OR Ireland[tiab] OR Irish[tiab] OR Ital*[tiab] OR Latvia*[tiab] OR Lithuania*[tiab] OR Luxemb*[tiab] OR Malt*[tiab] OR Netherlands[tiab] OR Dutch[tiab] OR Poland[tiab] OR Polish[tiab] OR Portug*[tiab] OR Romania*[tiab] OR Slovakia*[tiab] OR Slovenia*[tiab] OR Spain[tiab] OR Spanish[tiab] OR Sweden[tiab] OR Swedi*[tiab])

Additional papers:

<https://pubmed.ncbi.nlm.nih.gov/33547453/>

**Hierin vermeld je:**

onderzoeksvraag

zoekdatum

database(s)

aantal treffers/ gevonden records

zoekstrategie per aspect van de zoekvraag

kopie van de zoekgeschiedenis uit de database

In het logboek beschrijf je ook de afwegingen die je rondom jouw search maakt. Dit is zeker voor later handig: je weet waarom je welke keuzes hebt gemaakt. Daarnaast is dit een verantwoording van jouw zoekstrategie.

define the components of the search question

Each search question has four components remembered best by the mnemonic-PICO.9 1.Patient (P) What is the patient group of intereste.g. acute cholecystitis.

People living in the EU

 2.Intervention (I) What is the intervention of intereste.g. expectant management.

Trial hesitancy / trial confidence

 3.Comparison (C) What is the comparisonintervention of interest e.g. surgery.

 4.Outcome (O) What is the primary outcome e.g.survival.

A search is usually started with two of the four components, usually patient and intervention. Depending on the question and retrieval, the search canbe further focused by adding rest of the components of the question.

identify the study design that will answer the search question

Identify the study design that will answer the search question - The study design for most therapeutic interventions is likely to be a randomized double blind clinical trial (RCT). For clinical questions where several RCTs are reported in the literature, a systematic review or meta-analysis might be the most appropriate article to review. The optimal study design to assess prognosis and harm is likely to be a prospective cohort study.

3) identify the study type that will answer the search question.

Identify the study type that will answer the search question - Most clinical questions can be answered by one of the four recognized study types i.e. diagnosis,prognosis, therapy and harm. Several databases including PubMed provide appropriate search filters toretrieve citations with the particular study design.

Search query:

COVID*[tiab] AND (hesitancy*[tiab] OR confidence*[tiab]) AND participation*[tiab] AND trial*[tiab]

COVID*[tiab] AND (hesitancy*[tiab] OR confidence*[tiab]) AND willingness*[tiab] AND trial*[tiab]

COVID*[tiab] AND willingness*[tiab] AND trial*[tiab]

COVID*[tiab] AND willingness*[tiab] AND trial*[tiab]

COVID*[tiab] AND hesitancy*[tiab] AND trial*[tiab]

vaccine*[tiab] AND hesitancy*[tiab] AND trial*[tiab]

vaccine*[tiab] AND willingness*[tiab] AND trial*[tiab]

**vaccin*[tiab] AND willing*[tiab] AND trial[tiab]**

**COVID*[tiab] AND (hesitancy*[tiab] OR confidence*[tiab]) AND trial*[tiab]**

- Specific hesitancy per target groups (underrepresented)
- Covid-19 coverage per country
- Covid-19 coverage per target groups

A systematic search of MEDLINE, Embase, PubMed, medRxiv, and bioRxiv

**Objectives:** Pregnant people are at increased risk of COVID-19 related morbidity and mortality, and vaccination presents an important strategy to prevent negative outcomes. However, pregnant people were not included in vaccine trials, and there is limited data on COVID-19 vaccines during pregnancy. The objectives of this systematic review were to identify the safety, immunogenicity, effectiveness, and acceptance of COVID-19 vaccination among pregnant people in the U.S.

**Data sources:** Four databases (PubMed, Web of Science, CINAHL, and Google Scholar) were used to identify eligible studies published from January 01, 2020, through February 06, 2022.

**Study eligibility criteria:** Inclusion criteria were peer-reviewed empirical research conducted in the U.S., published in English, and addressed one of the following topics: safety, immunogenicity, effectiveness, and acceptance of COVID-19 vaccination among pregnant people.

**Study appraisal and synthesis methods:** A narrative synthesis approach was used to synthesize findings. Critical appraisal was done using the Joanna Briggs Institute (JBI) tool.

**Results:** Thirty-two studies were identified. The majority of studies (n = 25) reported the use of Pfizer and Moderna COVID-19 vaccines among pregnant people; only six reported the Janssen vaccine. Of the 32 studies, 11 examined COVID-19 vaccine safety, 10 investigated immunogenicity and effectiveness, and 11 assessed vaccine acceptance among pregnant people. Injection site pain and fatigue were the most common adverse events. One case study reported immune thrombocytopenia (ITP). COVID-19 vaccination did not increase the risk of adverse pregnancy or neonatal outcomes in comparison to unvaccinated pregnant people. After COVID-19 vaccination, pregnant people elicited a robust immune response, and vaccinations conferred protective immunity to newborns through breast milk and the placental transfer. COVID-19 vaccine acceptance was low among pregnant people in the U.S. African American race, Hispanic ethnicity, younger age, low education, prior refusal of the influenza vaccine, and lack of provider counseling were associated with low vaccine acceptance.

**Conclusions:** Peer-reviewed studies support COVID-19 vaccine safety and protective effects on pregnant people and their newborns. Future studies that use rigorous methodologies and include diverse populations are needed to confirm current findings. In addition, targeted and tailored strategies are needed to improve vaccine acceptance especially among minorities.
